# The Impact of Virtual Reality Training on Immune-Related Mechanisms in Alzheimer’s Disease

**DOI:** 10.1101/2025.02.22.25322714

**Authors:** Nazuke Yusufu, Li Hongyan

## Abstract

**Objective:** There is currently lack of research on the molecular mechanisms of VR therapy. In this study, we aimed to analyze the differential gene expression profiles and functional pathway enrichment of peripheral blood samples of AD related Mild Cognitive Impairment (MCI)patients before and after VR treatment using high-throughput sequencing methods and compare to normal controls to explore the targeted genes and potential intervention mechanisms of VR treatment.

**Methods:** Select five AD patients and five normal controls, collect peripheral blood samples, perform whole transcriptome sequencing to screen for differentially expressed mRNA genes before and after VR treatment and in comparison with the control group. Combine with GO and KEGG pathway enhancement to elucidate the underlying biological mechanisms and related pathways to predict the targeted genes and potential intervention mechanisms of VR treatment.

**Results:** Pre- and post-VR treatment group comparisons revealed a total of 1167 significantly differentially expressed genes in the mRNA data, with 165 genes upregulated and 1002 downregulated. Among these, ZFP36, FOS, JUN, FOSB, and PMAIP1 were identified as disease-related genes. Furthermore, immune infiltration analysis indicated that PMAIP1, ZFP36, and FOSB were correlated with immune cells. GO analysis highlighted significant enrichment in immune-related pathways.

**Conclusion:** VR treatment may modulate immune and transcriptional regulatory mechanisms, potentially contributing to its therapeutic efficacy in AD. The specific mechanisms require further research and validation.

## 1. Introduction

Alzheimer’s Disease (AD) is a neurodegenerative disorder that impairs intellectual development and disrupts neurocognitive functions ^[1]^. Currently, the etiological theories and research on AD mainly focus on areas such as amyloid protein deposition, hyperphosphorylation of tau protein, immune inflammation, vascular factors, oxidative stress, mitochondrial dysfunction, cholinergic neuronal abnormalities, protein misfolding, TDP43, alpha-synuclein, intestinal microbiota dysbiosis, and programmed cell death,etc. Therefore, if research on the therapeutic mechanisms related to AD starts from the aforementioned perspectives and combines with the pathogenesis, it can provide a scientific basis for finding new therapeutic targets and exploring molecular mechanisms for AD.

In cognitive rehabilitation therapy for AD, Virtual Reality (VR) is an important non-pharmacological intervention. Studies have shown that VR can improve the overall cognitive function, executive function, and emotional state of patients with cognitive impairment ^[2-4]^. However, there is currently lack of research on the molecular mechanisms of VR therapy. In this study, we aimed to analyze the differential gene expression profiles and functional pathway enrichment of peripheral blood samples of AD related Mild Cognitive Impairment (MCI)patients before and after VR treatment using high-throughput sequencing methods and compare to normal controls to explore the targeted genes and potential intervention mechanisms of VR treatment.

## 2. Materials and Methods

### 2.1. Study subjects and ethical considerations

Participants were selected from the Neurology Department of People’s Hospital of Xinjiang between January 2023 and January 2024. The average age was 62.5±8.2 years (age range 55-80). Patients were excluded if they had any of the following: 1. advanced cancer, stroke, active epilepsy, depression, or a history of mental illness; 2. disease that significantly affects cognitive assessment (e.g. hypothyroidism); 3. Impairments in hearing, vision, physical activity, or any other communication problems that may impact test completion; 4. Exclude patients who are taking medications for cognitive enhancement or other specialized drugs. A total of 10 participants were included in analysis, divided into 2 groups: normal control (NC, n=5) and AD -related MCI (n=5).

The enrollment criteria for NC were: No or mild memory impairment; no objective cognitive impairment; normal range in Montreal Cognitive Assessment (MoCA)?Mini-Mental State Examination (MMSE) score; Clinical dementia rating (CDR)=0; Activity of daily living scale(ADL) is normal.

Enrollment for AD patients were: Meets the diagnostic criteria for AD-related MCI as outlined in the NIA-AA 2011 guidelines^[5]^; Able to cooperate in completing cranial MRI, blood, and/or cerebrospinal fluid examinations; In this study, MoCA and MMSE scores are above the threshold for dementia; CDR scale score equals 0.5; ADL scale score is less than 25 points (indicating slight impairment or normal instrumental activities of daily living).

This study was approved by the Xinjiang People’s Hospital Ethics Committee in China (Approval Number: KY2024052445). It conformed to the Declaration of Helsinki, and written informed consent was obtained from all participants or their next of kin.

### 2.2. Methods

AD -related MCI group receives cognitive rehabilitation treatment based on immersive VR technology, three times a week for 20-30minutes per session, for a duration of 8 weeks. collect peripheral blood samples, perform whole transcriptome sequencing to screen for differentially expressed mRNA genes before and after VR treatment and in comparison with the control group. Combine with GO and KEGG pathway enhancement to elucidate the underlying biological mechanisms to predict the targeted genes and potential intervention mechanisms of VR treatment.

The R Bioconductor package DESeq2 was utilized to screen out the differentially expressed genes (DEGs). The P value for correction <0.05 and fold change>2 or < 0.5 were set as the cut-off criteria for identifying DEGs. To sort out functional categories of DEGs, Gene Ontology (GO) terms and KEGG pathways were identified using KOBAS 2.0 server Hyper geometric test and Benjamín-Hochberg FDR controlling procedure were used to define the enrichment of each term.

## 3. Results

### 3.1 Transcriptome analysis of differentially expressed genes in before VR treatment for AD(ADB), after VR treatment for AD (ADA) and normal controls(C)

ADA_vs_ADB comparisons revealed a total of 1167 significantly differentially expressed genes in the mRNA data, with 165 genes upregulated and 1002 downregulated. Gene Ontology (GO) functional enrichment analysis was performed on the genes exhibiting significant differential expression, Upregulated genes are predominantly enriched in pathways related to immune response, inflammatory response; downregulated genes are mainly concentrated in pathways associated with inflammatory response, endoplasmic reticulum stress response, and related processes. Fig 1

**Fig 1.**
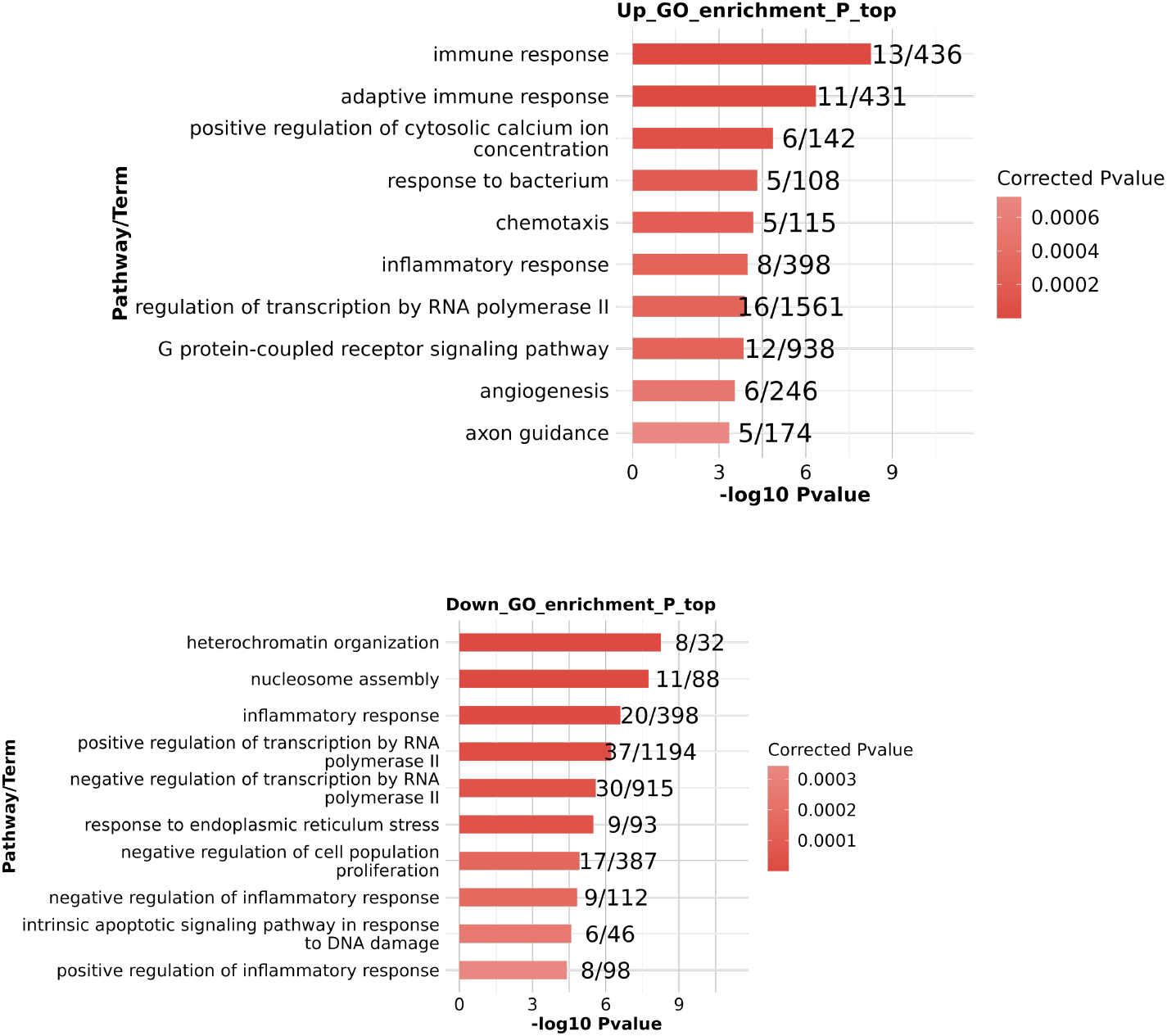
GO analysis of DEGs ADA_vs_ADB.

In the ADA_vs_C group, a total of 78 differentially expressed genes exhibited significant differential expression, among which 4 were upregulated and 74 were downregulated. The downregulated genes were primarily enriched in signaling pathways such as inflammatory response. Fig 2

**Fig 2.**
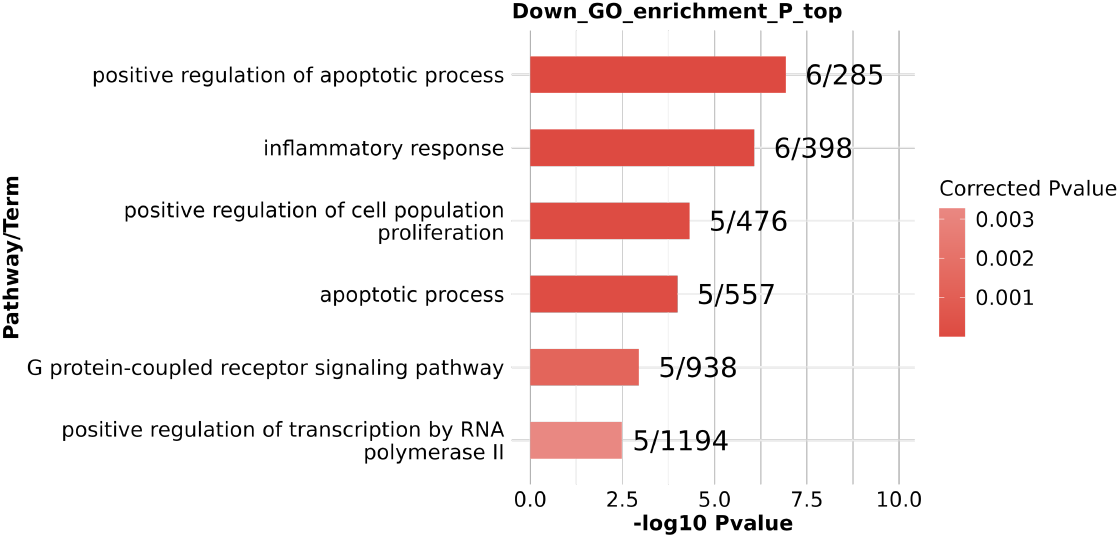
GO analysis of DEGs ADA_vs_C.

In the ADB_vs_C group, a total of 607 differentially expressed genes exhibited significant differential expression, with 421 genes upregulated and 186 genes downregulated. The upregulated genes were predominantly enriched in signaling pathways such as inflammatory response, while the downregulated genes were mainly concentrated in signaling pathways including chemokine-mediated signaling pathway, inflammatory response, and adaptive immune response. Fig 3

**Fig 3.**
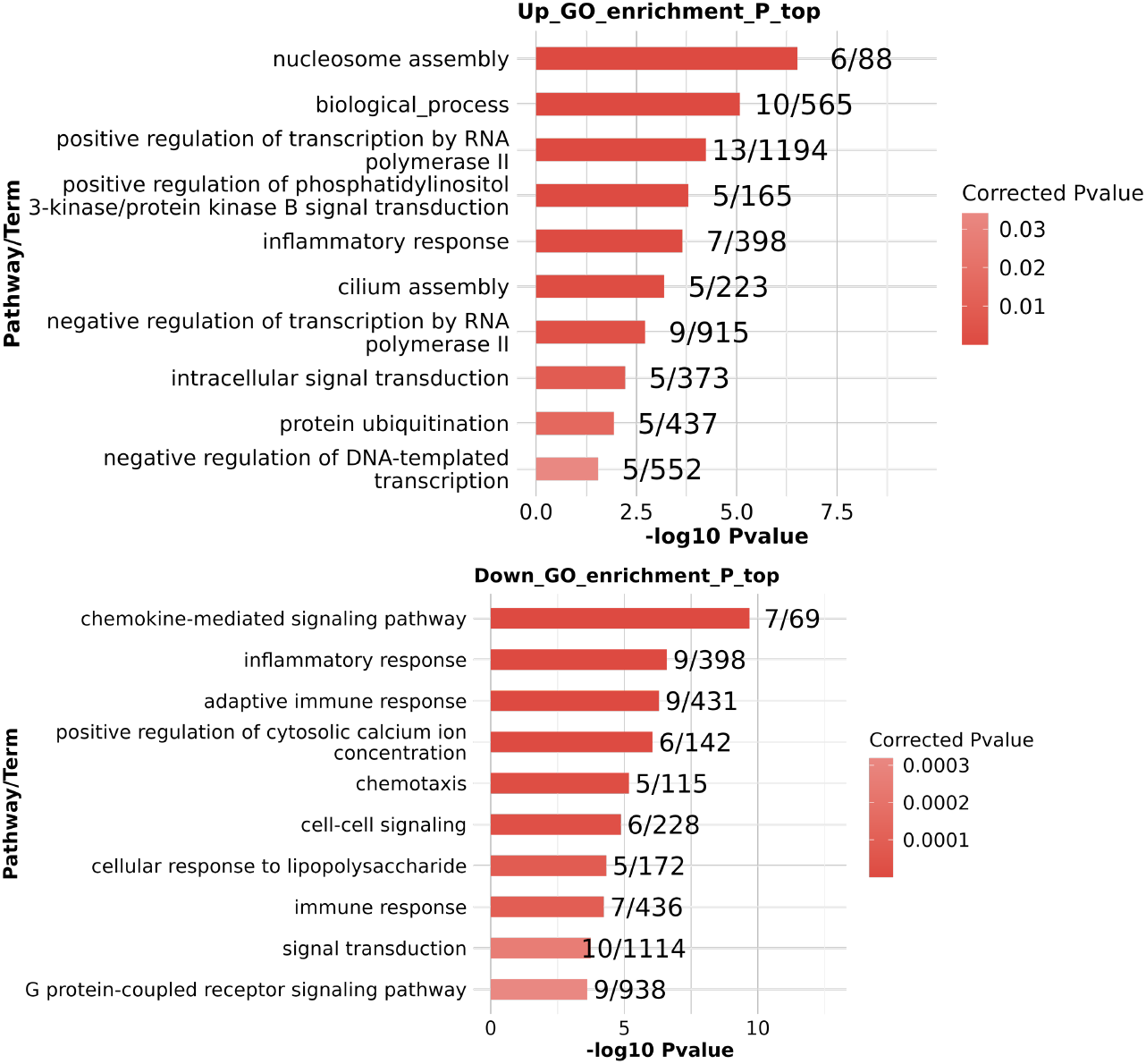
GO analysis of DEGs ADB_vs_C.

### 3.2 Dynamic analysis of DEGs

Based on transcriptomic data, we employed K-means clustering to perform differential gene expression trend analysis and categorized the genes into nine functional clusters. Notably, the DEGs within cluster 4 exhibited a relatively consistent trend of expression changes across samples in each group, which may be associated with the therapeutic effects achieved through virtual reality technology intervention. Fig 4

**Fig 4.**
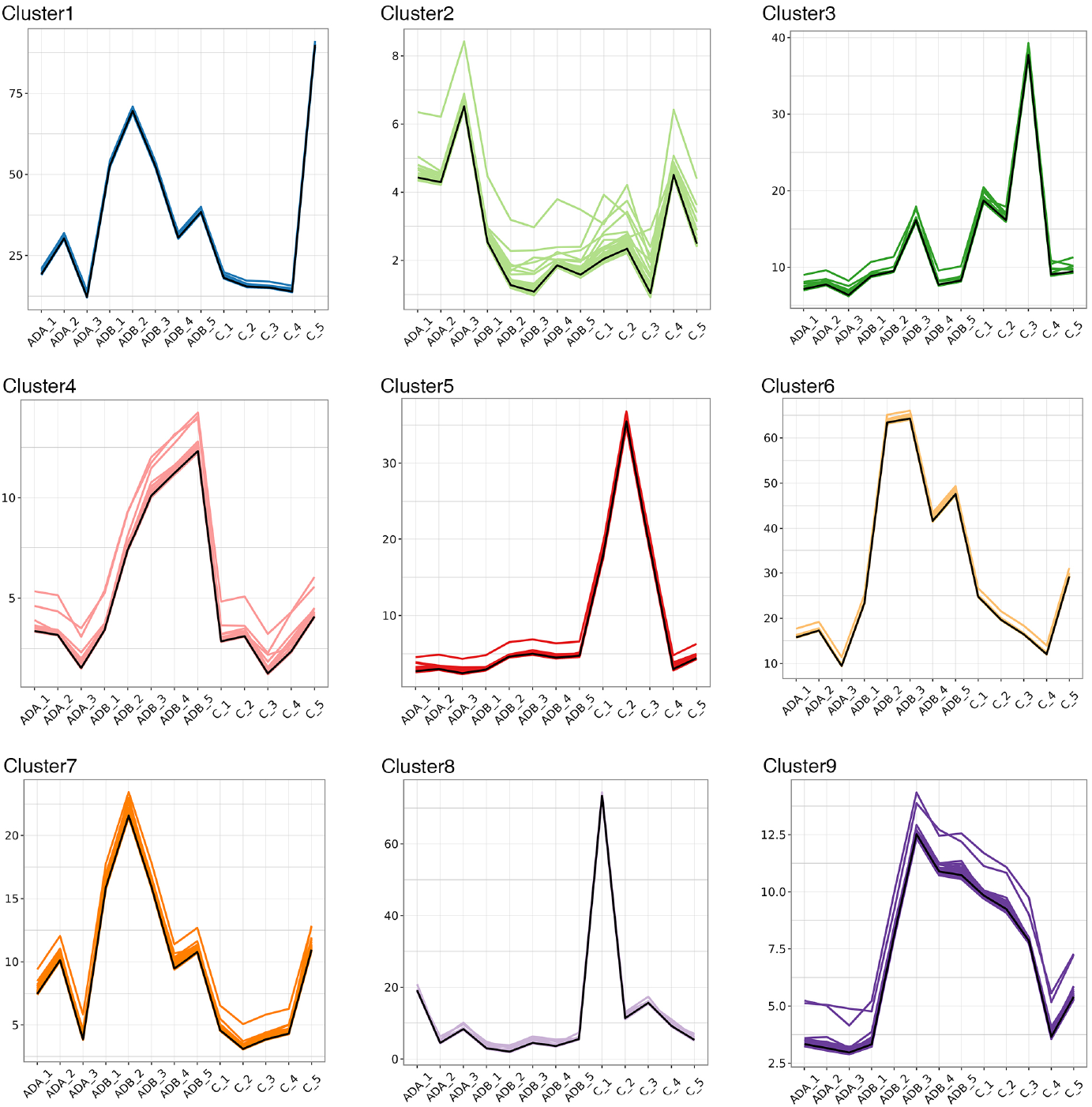
K-means clustering DEGs trend analysis.

GO enrichment analysis revealed that the genes in this cluster were primarily enriched in pathways such as nucleosome assembly, negative regulation of transcription by RNA polymerase II, biological processes, positive regulation of transcription by RNA polymerase II, heterochromatin formation, immune response, and regulation of transcription by RNA polymerase II. Fig 5

**Fig 5.**
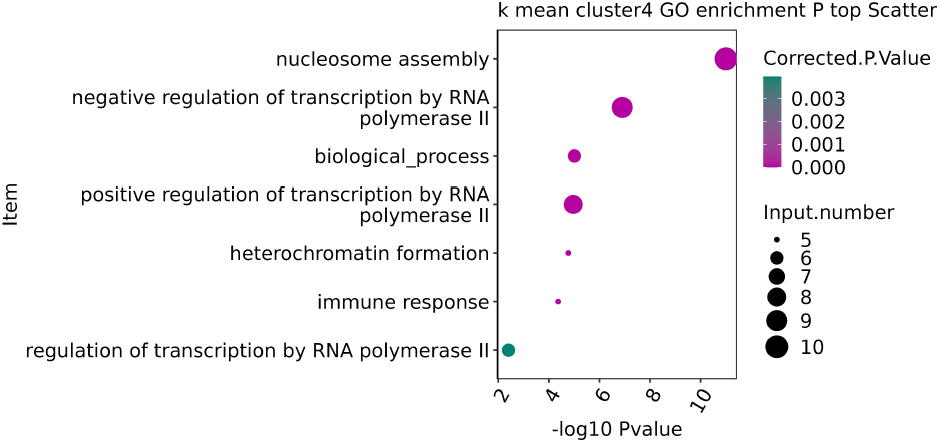
K-means cluster4 GO analysis.

Among these, ZFP36, FOS, JUN, FOSB, and PMAIP1 were identified as disease-related genes. Furthermore, immune infiltration analysis indicated that PMAIP1, ZFP36, and FOSB were correlated with immune cells. Given that the GO analysis highlighted significant enrichment in immune-related pathways, we further investigated the correlations between ZFP36, FOS, JUN, FOSB, PMAIP1, and immune cells (|cor| > 0.3 & p < 0.05). The analysis revealed that PMAIP1 exhibited a significant negative correlation with regulatory T cells (Tregs) and monocytes, while showing a significant positive correlation with neutrophils. Similarly, ZFP36 demonstrated a significant negative correlation with regulatory T cells and monocytes. Additionally, FOS was significantly negatively correlated with regulatory T cells (Tregs), activated NK cells, and monocytes, but significantly positively correlated with neutrophils. These findings suggest potential roles of these genes in modulating immune cell interactions and responses. Fig 6

**Fig 6.**
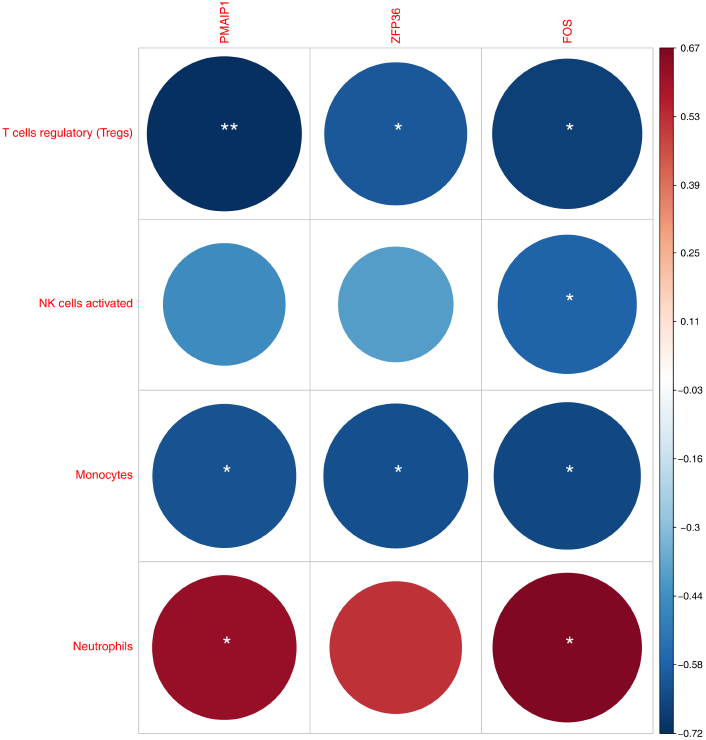
immune infiltration analysis.

## 4. Discussion

AD is the predominant form of dementia, presenting significant challenge globally. Its etiology is complex and multifactorial. Over the past few years, VR technology has emerged as a novel tool for the assessment and rehabilitation of AD. To elucidate the molecular mechanisms underlying VR-based treatment for AD, we obtained transcriptomic data through RNA sequencing (RNA-seq) from healthy controls, AD patients before and after VR treatment. Functional enrichment analysis using GO revealed that the majority of enriched pathways were associated with immune and inflammatory responses. Further analysis of DEGs using K-means clustering identified that the expression trends of DEGs in cluster 4 were relatively consistent across samples within each group, potentially correlating with the therapeutic effects of VR treatment. GO enrichment analysis of this cluster showed that the genes were primarily enriched in pathways related to immune response and regulation of transcription by RNA polymerase II. Notably, genes such as ZFP36, FOS, JUN, and FOSB, which are transcription factors, were prominently involved in these pathways.

ZFP36L1, a member of the ZFP36 family, has been strongly implicated in the regulation of autoimmunity ^[6]^. Study showed that ZFP36L1 was identified as a pathogenetic marker of AD, and a strong association between ZFP36L1 and the immune response was proposed ^[7]^. ZFP36L1 were the best co-diagnostic markers for AD and CKD based on the results of Least Absolute Shrinkage Selection Operator analysis and the random forest algorithm ^[8]^. The transcriptome sequencing further confirmed that the pathological symptoms of AD could be reversed by inhibiting the ERK/FOS axis to alleviate the inflammatory response ^[9]^. Activated c-Jun is present in neurofibrillary tangles in Alzheimer’s disease brains ^[10]^. Inhibition of c-Jun kinase provides neuroprotection in a model of Alzheimer’s disease ^[11]^. ΔFosB regulates gene expression and cognitive dysfunction in a mouse model of Alzheimer’s disease ^[12]^. Genome-wide profiling reveals functional diversification of ΔFosB gene targets in the hippocampus of an Alzheimer’s disease mouse model ^[13]^.

The treatment of AD may involve the restoration of dysregulated transcription factors, leading to the positive and negative feedback regulation of their target genes, particularly those related to immune responses. PMAIP1, a member of the BCL2 family, plays a role in the process of apoptosis ^[14]^. The therapeutic intervention in AD might modulate the expression of apoptosis-related genes, such as PMAIP1, thereby inhibiting neuronal cell death or senescence.

These findings suggest that VR treatment may modulate immune and transcriptional regulatory mechanisms, potentially contributing to its therapeutic efficacy in AD. The specific mechanisms require further research and validation.

## Data Availability

All data produced in the present study are available upon reasonable request to the authors

## Notes

### Competing Interest Statement

The authors have declared no competing interest.

### Funding Statement

This study did not receive any funding

### Author Declarations

This study was approved by the Xinjiang People's Hospital Ethics Committee in China (Approval Number: KY2024052445).

## References

1. Srivastava S, Ahmad R, Khare SK. Alzheimer’s disease and its treatment by different approaches: A review. European Journal of Medicinal Chemistry. 2021; 216:113320.

2. Stavropoulou I, Sakellari E, Barbouni A, Notara V. Community-based virtual reality interventions in older adults with dementia and/or cognitive impairment: A systematic review. Experimental Aging Research. 2024;40(2):123–140.

3. Bauer ACM, Andringa G. The potential of immersive virtual reality for cognitive training in elderly. Gerontology. 2020;66(4):503–513.

4. Moulaei K, Sharif H, Bahaadinbeigy K, et al. Efficacy of virtual reality-based training programs and games on the improvement of cognitive disorders in patients: A systematic review and meta-analysis. BMC Psychiatry. 2024;24(1):1–12.

5. Sperling R A, Aisen P S, Beckett L A, et al. Toward defining the preclinical stages of Alzheimer’s disease:recommendations from the National Institute on Aging-Alzheimer’s Association workgroups on diagnostic guidelines for Alzheimer’s disease[J]. Alzheimers Dement, 2011, 7(3):280–292.

6. Makita S, Takatori H, Nakajima H. Post-transcriptional regulation of immune responses and inflammatory diseases by RNA-binding ZFP36 family proteins. Frontiers in Immunology. 2021; 12:711633.

7. Tian Y, Lu Y, Cao Y, et al. Identification of diagnostic signatures associated with immune infiltration in Alzheimer’s disease by integrating bioinformatic analysis and machine-learning strategies. Frontiers in Aging Neuroscience. 2022; 14:919614.

8. Li J, Li Y, Niu J, et al. Exploration of the shared genetic biomarkers in Alzheimer’s disease and chronic kidney disease using integrated bioinformatics analysis. Medicine. 2023;102(44): e35555.

9. Wang C, Cui X, Dong Z, et al. Attenuated memory impairment and neuroinflammation in Alzheimer’s disease by aucubin via the inhibition of ERK-FOS axis. International Immunopharmacology. 2024; 126:111312.

10. Pearson AG, Byrne UTE, MacGibbon GA, et al. Activated c-Jun is present in neurofibrillary tangles in Alzheimer’s disease brains. Neuroscience Letters. 2006;398(3):246–250.

11. Braithwaite SP, Schmid RS, He DN, et al. Inhibition of c-Jun kinase provides neuroprotection in a model of Alzheimer’s disease. Neurobiology of Disease. 2010;39(3):311–317.

12. Corbett BF, You JC, Zhang X, et al. ΔFosB regulates gene expression and cognitive dysfunction in a mouse model of Alzheimer’s disease. Cell Reports. 2017;20(2):344–355.

13. You JC, Stephens GS, Fu CH, et al. Genome-wide profiling reveals functional diversification of ΔFosB gene targets in the hippocampus of an Alzheimer’s disease mouse model. PLoS One. 2018;13(2): e0192508.

14. Ling Y, Hu L, Chen J, et al. The mechanism of mitochondrial metabolic gene PMAIP1 involved in Alzheimer’s disease process based on bioinformatics analysis and experimental validation. Clinics. 2024; 79:100373.

